# Identification of potential novel combination antibiotic regimens based on drug-susceptibility and genetic diversity of Gram-negative bacteria causing neonatal sepsis in low- and middle-income countries

**DOI:** 10.1101/2023.10.20.23296805

**Authors:** Biljana Kakaraskoska Boceska, Tuba Vilken, Basil Britto Xavier, Christine Lammens, Sally Ellis, Seamus O’Brien, Renata Maria Augusto da Costa, Aislinn Cook, Neal Russell, Julia Bielicki, Eitan Naaman Berezin, Emmanual Roilides, Maia De Luca, Lorenza Romani, Daynia Ballot, Angela Dramowski, Jeannette Wadula, Sorasak Lochindarat, Suppawat Boonkasidecha, Flavia Namiiro, Hoang Thi Bich Ngoc, Tran Minh Dien, Tim R. Cressey, Kanchana Preedisripipat, James A. Berkley, Robert Musyimi, Charalampos Zarras, Trusha Nana, Andrew Whitelaw, Cely Barreto da Silva, Prenika Jaglal, Willy Ssengooba, Samir K. Saha, Mohammad Shahidul Islam, Marisa Marcia Mussi-Pinhata, Cristina Gardony Carvalheiro, Laura Piddock, Surbhi Malhotra-Kumar, Michael Sharland, Youri Glupczynski, Herman Goossens

## Abstract

**Objectives:** Several recent studies highlight the high prevalence of resistance to multiple antibiotic classes used in current treatment regimens for neonatal sepsis and new treatment options are urgently needed. We aimed to identify potential new combination antibiotic treatment regimens by investigating the drug-resistance and genetic profiles of the most frequently isolated Gram-negative bacteria causing neonatal sepsis in low- and middle-income countries (LMICs) in the NeoOBS study.

**Material and methods:** Gram-negative bacteria isolated from neonates with culture-confirmed sepsis from 13 clinical sites in nine countries, mainly LMICs, were analyzed. Culture-based identification was followed by whole-genome sequencing (WGS). Minimal inhibitory concentrations (MICs) for 8 antibiotics were determined for a representative subset of 108 isolates.

**Results:** Five bacterial species, *Klebsiella pneumoniae* (n=135), *Acinetobacter baumannii* (n=80), *Escherichia coli* (n=34), *Serratia marcescens* (n=33) and *Enterobacter cloacae* complex (ECC) (n=27) accounted for most Gram-negative bacterial isolates received (309/420, 74%). Extended-spectrum β-lactamases (ESBL) genes mostly belonging to CTX-M-15 were found in 107 (79%) *K. pneumoniae* isolates and 13 (38%) *E. coli*, as well as in 6 (18%) and 10 (37%) *S. marcescens* and ECC isolates, respectively. Carbapenem resistance genes were present in 41 (30%) *K. pneumoniae,* while 73 (91%) of *A. baumannii* isolates were predicted to be MDR based on carbapenem resistance genes. Apart from *A. baumannii,* in which two major pandemic lineages predominated, a wide genetic diversity occurred at the intraspecies level with different MDR clones occurring at the different sites. Phenotypic testing showed resistance to the WHO first- and second- line recommended treatment regimens: 74% of *K. pneumoniae* isolates were resistant to gentamicin and 85% to cefotaxime; *E. coli* isolates showed resistance to ampicillin, gentamicin and cefotaxime in 90%, 38% and 47%, respectively. For the novel antibiotic regimens involving different combinations of flomoxef, fosfomycin and amikacin, the overall predicted MIC-determined susceptibility for Enterobacterales isolates was 71% (n=77) to flomoxef-amikacin, 76% (n=82) to flomoxef-fosfomycin and 79% (n=85) to fosfomycin-amikacin combinations, compared to 31% and 22% isolates susceptible to ampicillin-gentamicin and cefotaxime, respectively. ESBL-producing Enterobacterales isolates were 100% susceptible both to flomoxef-fosfomycin and flomoxef-amikacin and 92% to fosfomycin-amikacin.

**Conclusion:** Enterobacterales carried multiple resistance genes to cephalosporins, carbapenems and aminoglycosides. ESBL-producing *K. pneumoniae* and *E. coli* isolates were highly susceptible to the three new antibiotic combination regimens planned to be evaluated in the currently recruiting GARDP-sponsored NeoSep1 trial.

## Introduction/Background

In 2019, more than 560,000 neonatal deaths were associated with bacterial antimicrobial resistance (AMR), including nearly 140,000 deaths directly attributable to bacterial AMR [1]. The World Health Organization (WHO) reports that over 80% of these sepsis deaths could be prevented if there was improved treatment and infection prevention [2].

Most of these cases occur in low- and middle-income countries (LMICs) [3]. Of the multiple large studies recently conducted in these countries [1, 4–6], one systematic review [7] and several single-site reports [8–10] have shown that Gram-negative bacteria (GNB), such as *Klebsiella* spp., *Escherichia coli* and *Acinetobacter baumannii*, are considered the main cause of neonatal sepsis in approximately 40% episodes. Furthermore, these studies have demonstrated that the empiric treatment of neonatal sepsis currently recommended by WHO, which includes a narrow-spectrum β-lactam antibiotic in combination with gentamicin as a first line regimen and a 3^rd^ generation cephalosporin as a second line regimen [11], is increasingly compromised by high drug resistance rates, particularly due to the high prevalence of ESBLs and of aminoglycoside modifying enzymes (AMEs). Findings from these recent studies extend data from several previous reports from LMICs where extremely high rates of resistance to amoxicillin (80%), gentamicin (60%) and third-generation cephalosporins (>80%) were observed [12–17].

The NeoOBS study [6, 18] was a prospective, multicenter, observational cohort study investigating the management of neonatal sepsis in several countries, aiming to inform and enhance the design of the currently recruiting Global Antibiotic Research & Development Partnership (GARDP) - sponsored NeoSep1 antibiotic trial (ISRCTN48721236). The trial is investigating new antibiotic regimens for the treatment of neonatal sepsis that have activity against priority neonatal bacterial pathogens and have the potential to be available for wide use in LMICs.

Three generic antibiotics, amikacin, flomoxef and fosfomycin were selected that met the criteria for consideration in the trial [19], and their potential to be used in novel combined empirical regimens was assessed by a dynamic hollow-fiber infection model (HFIM) and by checkerboard assays [20–22]. The three combinations, fosfomycin-amikacin, fosfomycin-flomoxef and flomoxef-amikacin exhibited synergistic interactions measured by both bactericidal killing and the prevention of emergence of resistance.

The NeoOBS microbiology sub-study aimed to determine the level of antibiotic susceptibility of MDR Gram-negative bacteria to currently used antibiotics and to the novel drug combinations included in the NeoSep1 trial.

## Material and Methods

### Study setting

A prospective observational clinical study of neonatal sepsis was conducted between 2018 and 2020 at 19 hospitals across 11 predominantly LMIC countries from five WHO regions (Africa, America, Europe, Southeast Asia, and Western Pacific) [6]. Hospitalized infants <60 days of age with an episode of clinically suspected sepsis were eligible for enrollment.

### Strain collection and characterization of isolates

The Laboratory of Medical Microbiology (LMM) at the University of Antwerp received bacterial isolates from blood and cerebrospinal fluid (CSF) samples of neonates with culture-confirmed sepsis from 13 participating sites in 9 countries (Supplementary Table 1). LMM did not obtain bacterial isolates from 3 sites from India and 3 from China. All the participating sites followed a well-established microbiological protocol for collection, storage and shipment of the isolates to the central laboratory [18]. At LMM, species identification was verified using Microflex LT MALDI-TOF MS (Bruker Daltonics) and the MALDI Biotyper IVD reference library (2021).

### Selection of the isolates for inclusion in this microbiology study

Identification at species level was confirmed by WGS for 723 out of 1051 bacterial isolates. Gram positive were 303 (Coagulase-negative staphylococci (n=254) and *Staphylococcus aureus*, n=49), and 420 were Gram-negative, belonging to 23 different species. Enterobacterales, comprising *Klebsiella pneumoniae* (n=135), *Escherichia coli* (n=33), *Serratia marcescens* (n=34) and *Enterobacter cloacae* complex (n=27), as well as *Acinetobacter baumannii* (n=80) as non-fermenters, were the most common pathogens, together accounting for nearly three-quarters of Gram-negative isolates. The focus of this study was limited to Gram-negative bacteria.

For all analyses, the patient’s first clinical isolate from blood or CSF was selected. In case of mixed infection, isolates belonging to different species were also included. To avoid analyzing replicates of the same bacterial clone, only isolates displaying different genetic profiles were selected for *in vitro* susceptibility testing. Isolates belonging to a given species for which less than 20 isolates were obtained were not further analyzed.

### DNA isolation and Whole Genome Sequencing (WGS)

Genomic DNA isolation was done using the MasterPure complete DNA and RNA purification kit (Epicentre, Madison, WI, USA). Library and sample preparation were performed using Nextera XT sample preparation kit (Illumina) and for sequencing MiSeq (Illumina Inc., USA) was used. The raw data of the sequenced strains were quality checked and trimmed using trimmomatic v0.4.2. The second analysis was performed using BacPipe v.1.2.6 [23]. Built-in tools and databases were used for *de novo* assembly SPAdes (v.3.11.0) and Prokka (v1.11.1) for annotation. Assembled contigs were quality checked using checkM [24]. MLST for *E. coli* [25], *K. pneumoniae* [26], *Enterobacter* spp. [27] and *Acinetobacter* spp. [28] were determined according to MLST schemes of organism-specific reference databases. For core genome multilocus sequence typing (cgMLST), a gene-by-gene approach was utilized by developing a custom scheme for the specific study, assessing allelic loci distances using ChewBBACA [29]. Clonal relatedness was defined as ≤10 allelic differences between isolates for *A. baumannii* [29] and <12 for *Klebsiella* spp. [30]. Trees were visualized using Grapetree [31]. For strains for which phenotypic MIC susceptibility testing was not performed, the presence of one or more resistance genes to the tested antibiotics was used to reflect resistance.

### *In vitro* susceptibility testing

Eighty-seven *K. pneumoniae* and 21 *E. coli* isolates representing different genetic profiles were tested against first- and second- line WHO-recommended regimens (ampicillin, gentamicin and cefotaxime), as well as piperacillin-tazobactam, meropenem and three antibiotics (flomoxef, amikacin and fosfomycin) that are under investigation as a potential new regimen in combination. The activity of fosfomycin was assessed by agar dilution method. All tests were performed according to the latest guidelines of the EUCAST susceptibility testing standards and interpretation criteria (https://www.eucast.org/clinical_breakpoints).

For flomoxef, in the absence of formal EUCAST/CLSI clinical breakpoint, we used an MIC susceptibility breakpoint of ≤1 mg/L, based on a large retrospective study in patients with bacteremia due to ESBL producers, which reported a more favorable clinical outcome in patients treated with flomoxef when isolate MICs were ≤1 mg/L compared to those with MICs ≥2 mg/L [32].

The susceptibility of the isolates to the proposed combination regimens was analyzed based on the novel combination breakpoint thresholds determined by HFIM model and checkerboard assays [20–22]. If flomoxef MIC values are between 1 and 32 mg/L, combination with fosfomycin or amikacin extends flomoxef’s spectrum of activity, but only when MIC of the associated drug is in the range of the corresponding MIC breakpoints, i.e. ≤32 mg/L for fosfomycin and ≤16 mg/L for amikacin [20, 21]. Success of fosfomycin-amikacin combination is predicted if the product of the two individual antibiotic MICs is ≤256 mg/L [R. da Costa, personal communication, August 31, 2023].

## Results

In total, 420 Gram-negative bacterial isolates were received. The five most common identified species were: *K. pneumoniae* (n=135), *A. baumannii* (n=80), *E. coli* (n=34), *S. marcescens* (n=33) and *Enterobacter cloacae* complex (ECC) (n=27). These 309 Gram-negative isolates were obtained from 295 patients. The remaining 111/420 GNB isolates belonged to 18 different bacterial species were represented by less than 20 isolates each. The distribution of the five major GNB isolates per sites is shown in Figure 1.

**Figure 1.**
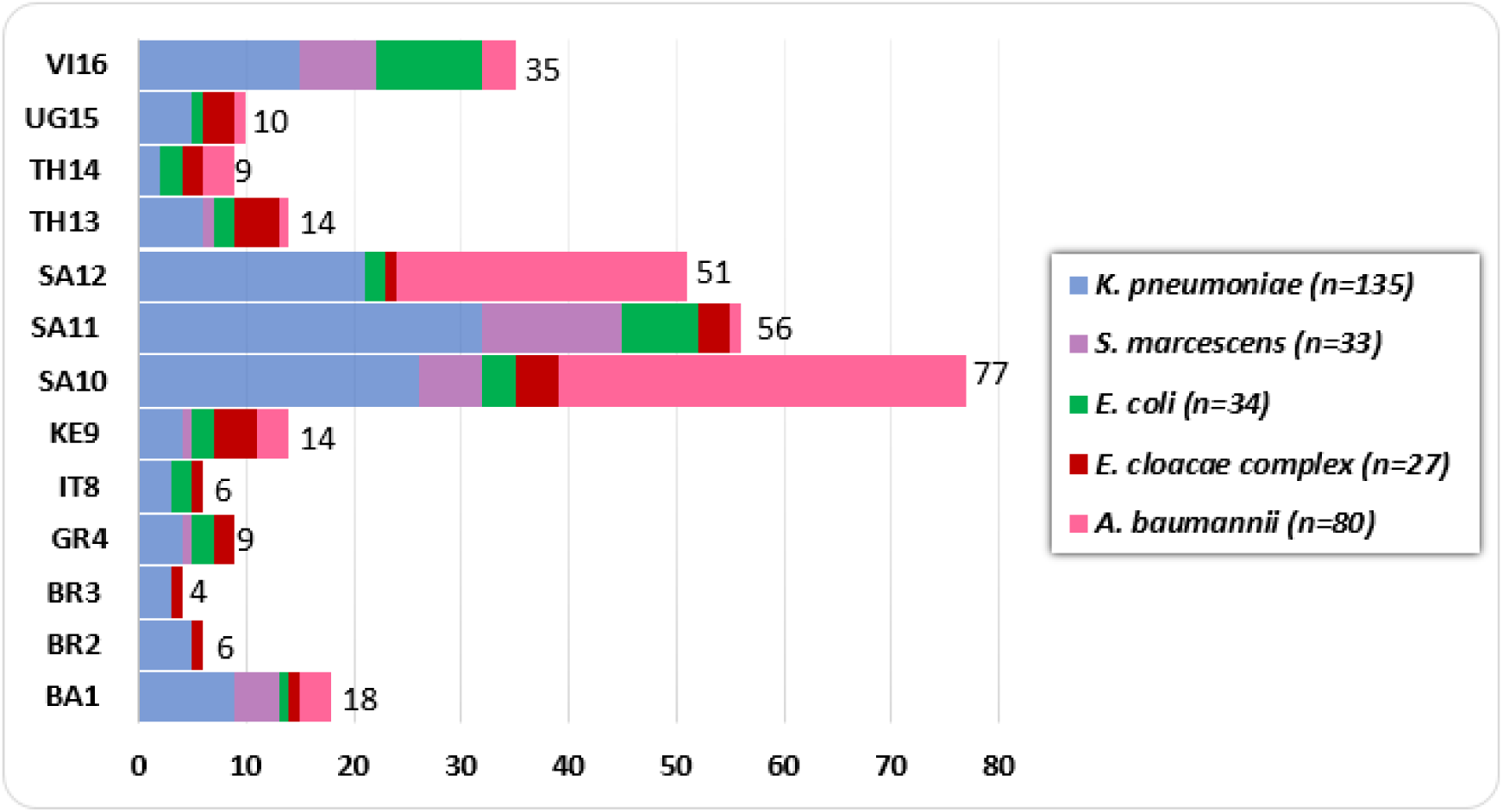
Distribution of the total number (indicated next to the bars) of the five most common GNB species analyzed by site. (n=309, one isolate per species per patient only, following removal of duplicates). (number of isolates correlates with the number of neonates, except for the following sites: TH13 (13 neonates - 14 isolates), SA12 (46 neonates - 51 isolates), SA11 (51 neonates - 56 isolates), SA10 (75 neonates - 77 isolates) and BR2 (5 neonates - 6 isolates)).

### Antibiotic susceptibility testing and resistance mechanisms

Ampicillin/gentamicin regimen had low rates of coverage, with 32% (34/108) of the isolates susceptible, and only 22% (24/108) susceptible to cefotaxime (Supplementary Table 2).

*K. pneumoniae* was the species with highest rates of resistance due to the high prevalence of ESBLs and of AMEs, found in 74% (64/87) and 63% (55/87) of the isolates, respectively. The rates of susceptibility to piperacillin-tazobactam differed between *K. pneumoniae* (35%) and *E. coli* (67%) due to the frequent occurrence of the *bla*_OXA-1_ gene among *K. pneumoniae* isolates. Carbapenem resistance was almost exclusive *to K. pneumoniae*, with 30% (26/87) strains resistant to meropenem versus 5% (1/21) of *E. coli*. The most prevalent carbapenem resistance genes were NDM-like (81%; n=21/26), followed by OXA-48-like (11%; n=3/26).

Susceptibility rates to flomoxef (71%) and amikacin (70%) were comparable to those observed for meropenem (75%) in the subset of 108 isolates with available MIC results. Flomoxef displayed good activity against ESBL-producing Enterobacterales (ESBL-PE) and resistance to this agent was due to class C β-lactamases and carbapenemases. Among the aminoglycosides, amikacin showed the best activity against ESBL-PE, since it was not affected by common AMEs (AAC(3)-II, ANT(2’)-I), which are known to modify gentamicin. Fosfomycin showed the strongest antibacterial activity with 90% of the phenotypically tested isolates being susceptible to this antibiotic. An almost perfect match (106/108) was found between susceptibility and resistance phenotypes and genotypes for all beta-lactams, including broad-spectrum penicillins, cephalosporins and carbapenem drugs, as well as for aminoglycosides. All ESBLs, AmpC and carbapenem resistance genes detected by genotyping correctly predicted strain susceptibility patterns, except for one *K. pneumoniae* isolate carrying the *bla*_NDM-1_ gene but found to be susceptible to meropenem (MIC of 0.06 mg/L). For aminoglycosides, all strains carrying resistance genes coding for modifying enzymes to gentamicin and/or to amikacin were confirmed as resistant to these drugs by MIC testing. All strains coding for 16S rRNA methylase genes displayed high level resistance to gentamicin and to amikacin (MIC > 256 mg/L for both agents). One *K. pneumoniae* isolate displayed high-level resistance to amikacin (MIC >256 mg/L) but did not contain a resistance gene that accounts for resistance to amikacin. Among the 11 fosfomycin-resistant strains, only two carried the gene encoding the fosfomycin-modifying enzyme (*fosA3* in one *E. coli* isolate with a fosfomycin of MIC >512 mg/L and *fosA5* in one *K. pneumoniae* isolate with an MIC of 64 mg/L). Besides *fosA3* and *fosA5*, genes coding other fosfomycin-modifying enzymes (*fosB*, *fosC*, *fosX*) were not found in the resistant strains. Also, no mutations of cell-wall transport systems (*glpT* and *uhpT*), their respective regulatory genes (*cyaA* and *ptsI*) nor *murA* targets were present in any of these strains.

The rates of susceptibility to the proposed new regimens were assessed using the novel combination breakpoint thresholds determined by HFIM assays [20–22]. For Enterobacterales isolates for which MIC determination was performed, prediction of susceptibility to at least one antibiotic combination was 77/108 (71%) isolates for flomoxef-amikacin, 85/108 (79%) for fosfomycin-amikacin and 82/108 (76%) for flomoxef-fosfomycin compared to 34/108 (31%) for ampicillin-gentamicin and 24/108 (22%) for cefotaxime (Table 1 and Supplementary Table 2). The three new combinations exhibited strong activity against ESBL-producing *E. coli* and *K. pneumoniae* isolates (excluding those strains producing an AmpC cephalosporinase and carbapenemase in addition to ESBL) and an excellent coverage with flomoxef-amikacin and flomoxef-fosfomycin of 100% (n=52/52) and 92% (n=48/52) with fosfomycin-amikacin combinations.

**Table 1.** Susceptibility of 108 Enterobacterales isolates to different combination regimens based on the novel combination breakpoint thresholds. [20–22]

| Enterobacterales* isolates tested for MIC determination | FOS<sup>°</sup>/FLX<sup>°</sup> | FOS/AMK<sup>°</sup> | FLX/AMK |
| --- | --- | --- | --- |
|  | N (% coverage) |  |  |
| **Cefotaxime-S (n=24)** | 24 (100) | 23 (96) | 24(100) |
| *ESBL-neg Enterobacterales* |  |  |  |
| **Cefotaxime-R/meropenem-S (n=58)** | 55 (95) | 52 (90) | 53 (91) |
| *ESBL-pos/Carba-Neg Enterobacterales (n=52)* | 52 (100) | 48 (92) | 52 (100) |
| *AmpC-pos/Carba-Neg Enterobacterales (n=6)* | 3 (50) | 4 (67) | 1 (17) |
| **Meropenem-R (n=26)** | 3 (12) | 10 (38) | 0 (0) |
| *Carba-Pos Enterobacterales\*\** |  |  |  |
| **Total number of isolates (n=108)** | 82 (76) | 85 (79) | 77 (71) |

\* *Klebsiella pneumoniae* (n=87), *Escherichia coli* (n=21)
<sup>°</sup> FOS: fosfomycin, FLX: flomoxef, AMK: amikacin
\*\* Among the 26 meropenem-resistant, carbapenemase-producing Enterobacterales isolates, 21 were concomitantly ESBL-producing and 5 were ESBL-negative.

Susceptibility of *S. marcescens* and ECC was predicted by the presence of resistance genes. Both species harbored genes coding for inducible chromosomal AmpC and displayed resistance to cefotaxime, as well as to flomoxef alone. In addition, genes coding for ESBLs were observed in 6/33 (18%) of *S. marcescens* and 10/27 (37%) of ECC isolates. Resistance to gentamicin was predicted in 4/33 (12%) of *S. marcescens* and in 10/27 (37%) of ECC.

The activity of amikacin was predicted to cover 91% of *S. marcescens* and 96% of ECC isolates, since genes coding for AMEs or for 16SrRNA methylase genes that confer resistance to this antibiotic were not widespread. Carbapenem resistance was predicted to be low, as only one isolate per species was found to carry an NDM-like gene.

Among *A. baumannii* isolates, 91% (73/80) harbored OXA-like carbapenemase genes (mostly OXA-23 and OXA-58) and 45% (36/80) co-harbored NDM-1 always in association with OXA-23, as well as resistance genes to several other classes of antimicrobials, including the 16S rRNA methylase genes, that confers high-level resistance to all aminoglycosides for clinical use.

### Characterization of Klebsiella pneumoniae

*K. pneumoniae* isolates were found in all 13 sites, and WGS analysis revealed high genetic diversity with a total of 56 different sequence types (STs) found among the 135 isolates obtained. Fifty-seven isolates belonging to 12 STs were predicted to be MDR, based on the presence of acquired drug resistance genes to 3 or more classes of antimicrobial agents. These MDR *K. pneumoniae* clones were isolated at all sites but with different geographic distribution of STs (Table 2). For instance, the predominant STs in South African sites were ST39, ST17 and ST14, while ST15 and ST147 were found in Vietnam and Bangladesh, respectively. As illustrated in Table 2, *K. pneumoniae* belonging to different STs were often co-circulating at different sites.

**Table 2.**
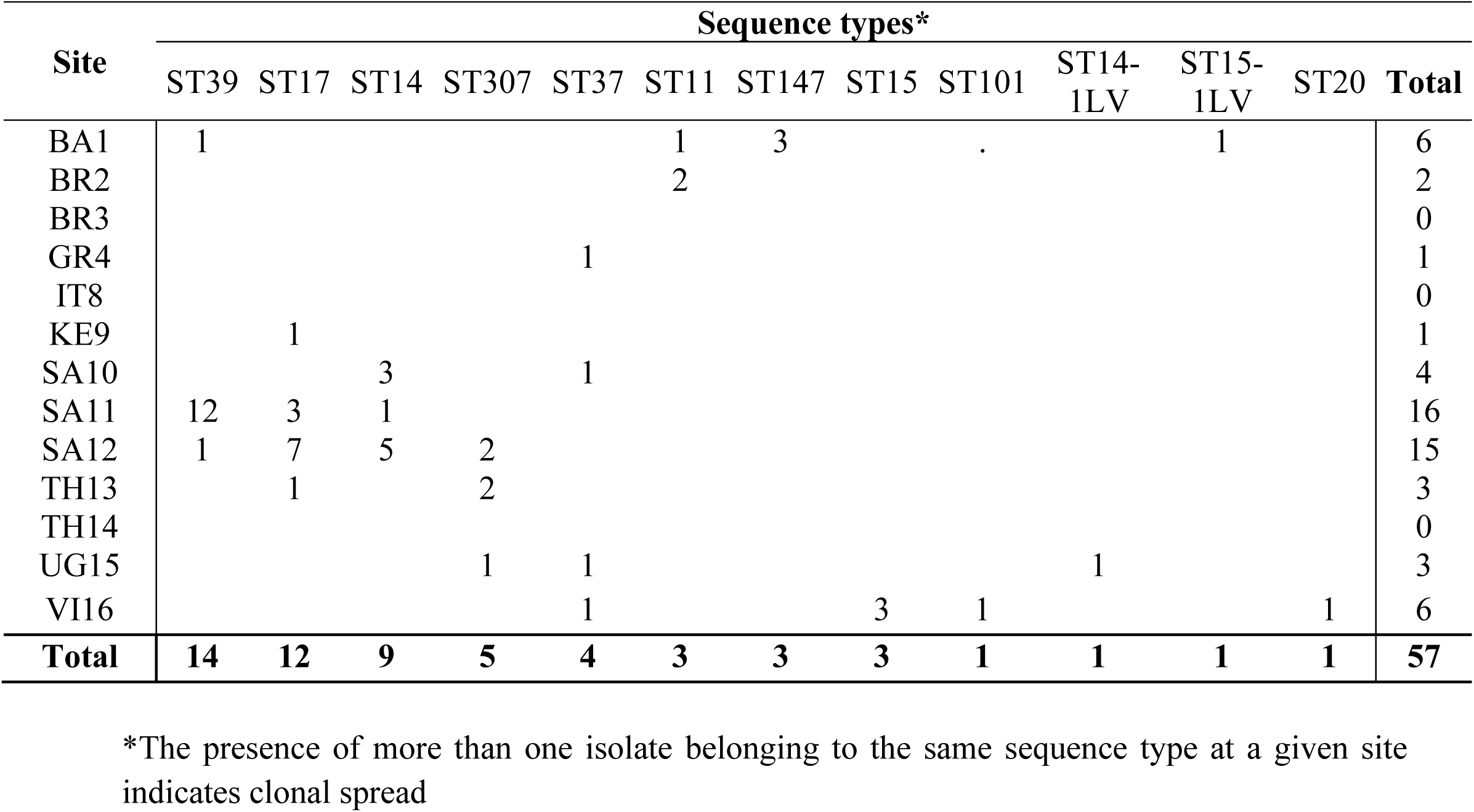
Distribution of the 12 most prevalent MDR STs of *K. pneumoniae* by site.

CgMLST revealed that isolates of the same ST found at different sites could be delineated in different clones (Figure 2). This analysis had, as expected, a much higher discriminatory power than classical multilocus sequence typing and it allowed the detection of distinct clones within a single ST, most notably for ST39, ST14 and ST17.

**Figure 2.**
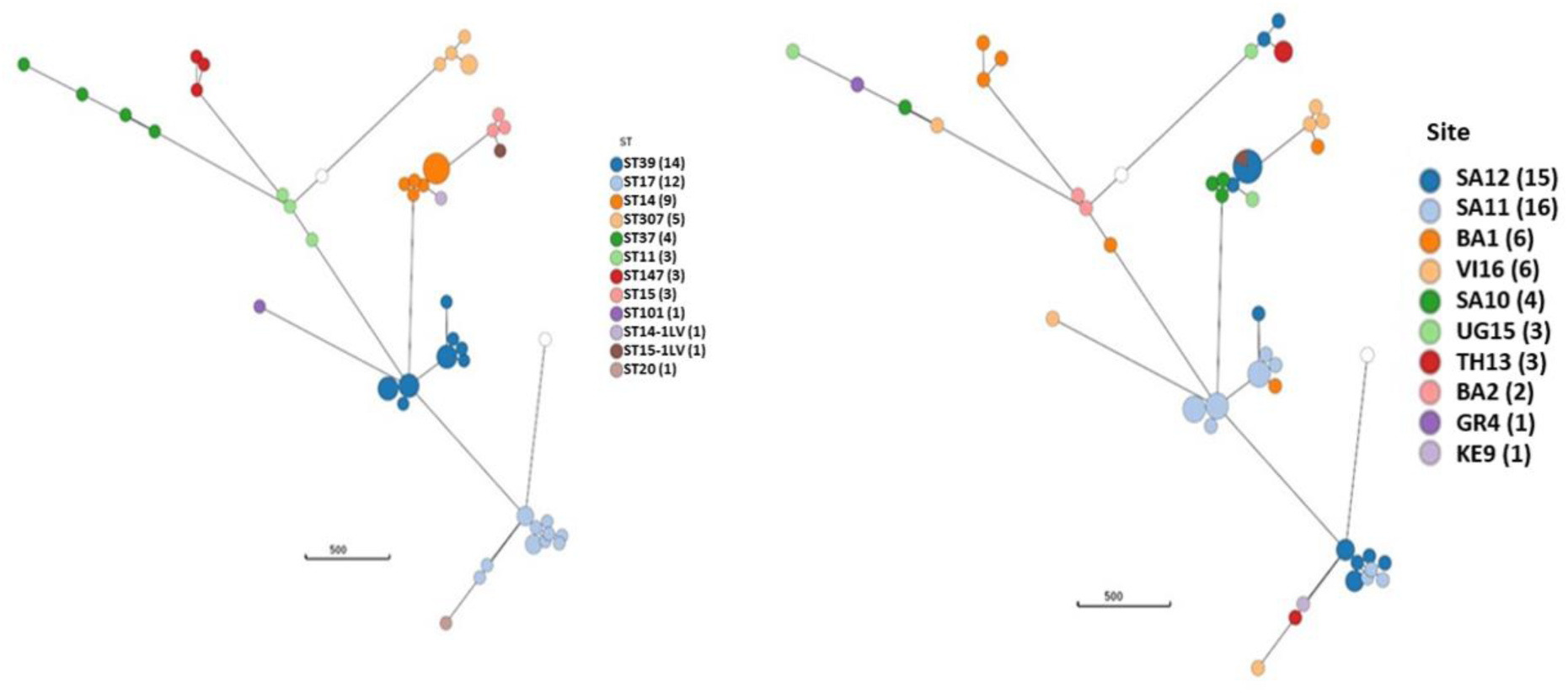
Minimum spanning tree of cgMLST analysis of MDR clones of *K. pneumoniae* (n=57) showing specific local site clustering. Left: ST based clustering Right: Site based clustering. The white circles represent public references.

In total, 83% (112/135) of *K. pneumoniae* isolates carried one or several ESBL genes encoding resistance to extended spectrum cephalosporins; this was confirmed phenotypically for 65/87 isolates (44 ESBL only and 21 in association with carbapenamase) tested phenotypically. ESBL coding genes were widely distributed and found in isolates from 12/13 sites; the *bla*_CTX-M-15_ gene was the most prevalent ESBL, present in 96 isolates at 10 sites. Other less frequently found ESBL genes were *bla* _CTX-M-14_ (in 7 isolates), *bla* _CTX-M-27_ (in 2 isolates) both at site VI16 in Vietnam, *bla*_SHV_ (in 5 isolates) and *bla*_TEM_ (in 2 isolates).

On the contrary, only six isolates carried AmpC genes, *bla*_MOX-2_ (n=3, site GR4) and *bla*_DHA-1_ (n=3 from BA1, TH13 and VI16).

All strains with carbapenemase coding genes were phenotypically resistant. These strains were found at 7 sites across five countries (Figure 3). The most frequent carbapenem resistance gene was *bla*_NDM_ being identified in 31/135 (23%) of *K. pneumoniae* isolates. Among the different variants, *bla*_NDM-1_ was the most prevalent (n=20) but other alleles (*bla*_NDM-4_ and *bla*_NDM-5_) were also found. As known from the epidemiology of carbapenemase producers, specific carbapenem resistance genes were associated with the geographic areas in which the isolates were found. Despite the small number of isolates, KPC producing strains were mostly found in Brazil, those carrying OXA-48-like carbapenemase in Bangladesh (BA1) and South Africa (SA11) and those with NDM-4 and NDM-5 in South-East Asia (VI16) and Asia (BA1) (Figure 3).

**Figure 3.**
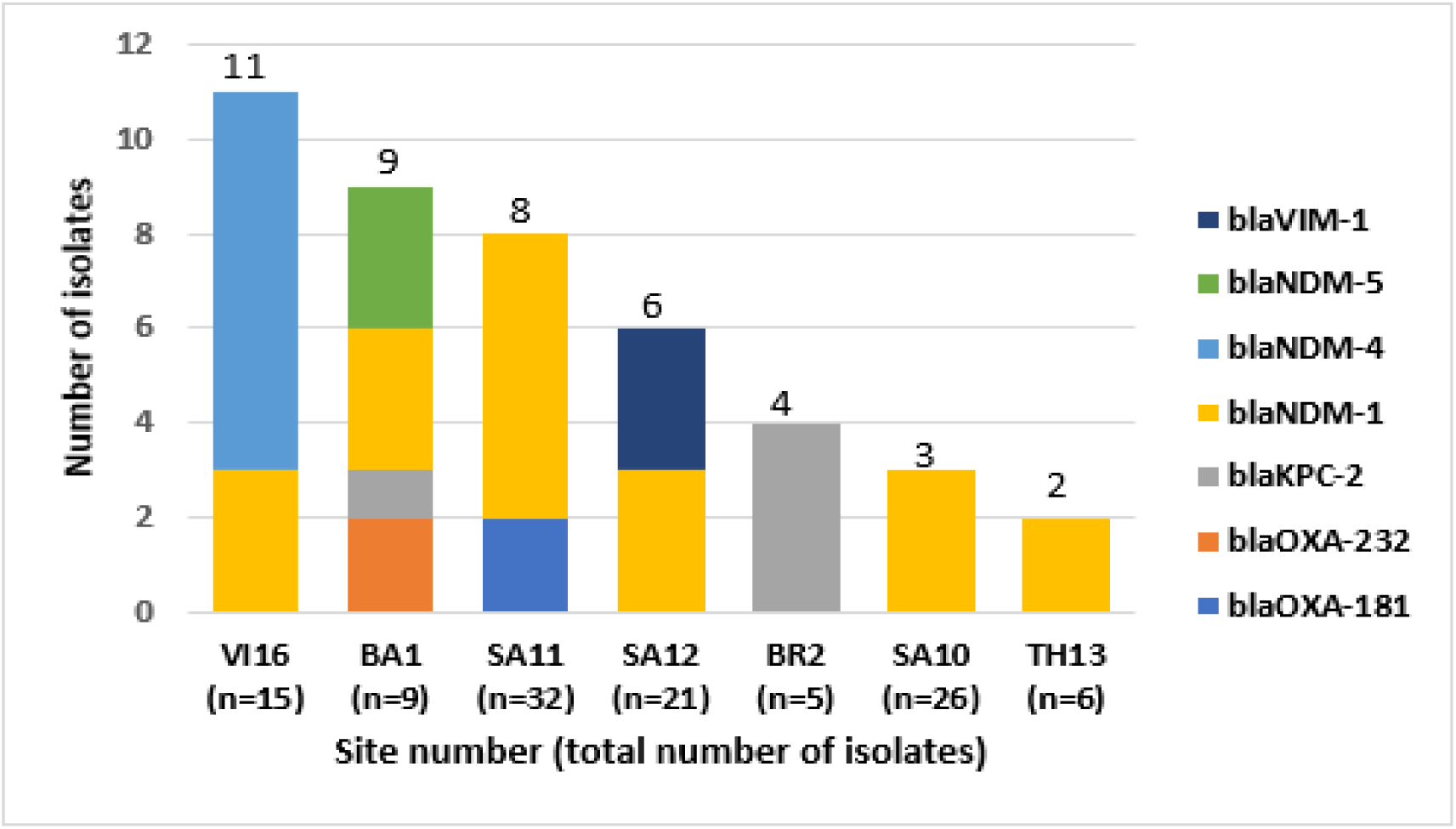
Distribution of carbapenem resistance genes of *K. pneumoniae* isolates (n=135) by site. (n) = total number of *K. pneumoniae* isolates per site. Sites that collected *K. pneumoniae* isolates lacking any carbapenem resistance genes are not shown in this figure (UG15 n=5, GR4 n=4, KE9 n=4, IT8 n=3, BR3 n=3 and TH14 n=2).

Acquired aminoglycoside resistance genes were found in 122/135 (90%) *K. pneumoniae* isolates (Figure 4), usually in association with ESBL- and/or with carbapenemase-coding genes. In particular the *aac(3)-II* genes which confers resistance to gentamicin, were frequently present (n=90; 67%), most often in association with *bla*_CTX-M-15_ ESBL gene (n=78). Other genes lIke *aph(3’)-VI* that are known to be associated with resistance to amikacin were less frequent and found only in 6 isolates at three sites, BA1, GR4 and IT8 (n=2 each). Genes coding for 16S rRNA methylases (*armA, rmtB, rmtC, rmtF*), which are known to confer resistance to all clinically used aminoglycosides, were identified in 23 isolates from 5 sites (BA1, SA10, SA11 SA12 and VI16) (Figure 4). The 16S rRNA methylase genes were carried only by carbapenemase-producing isolates, mostly NDM-producers.

**Figure 4.**
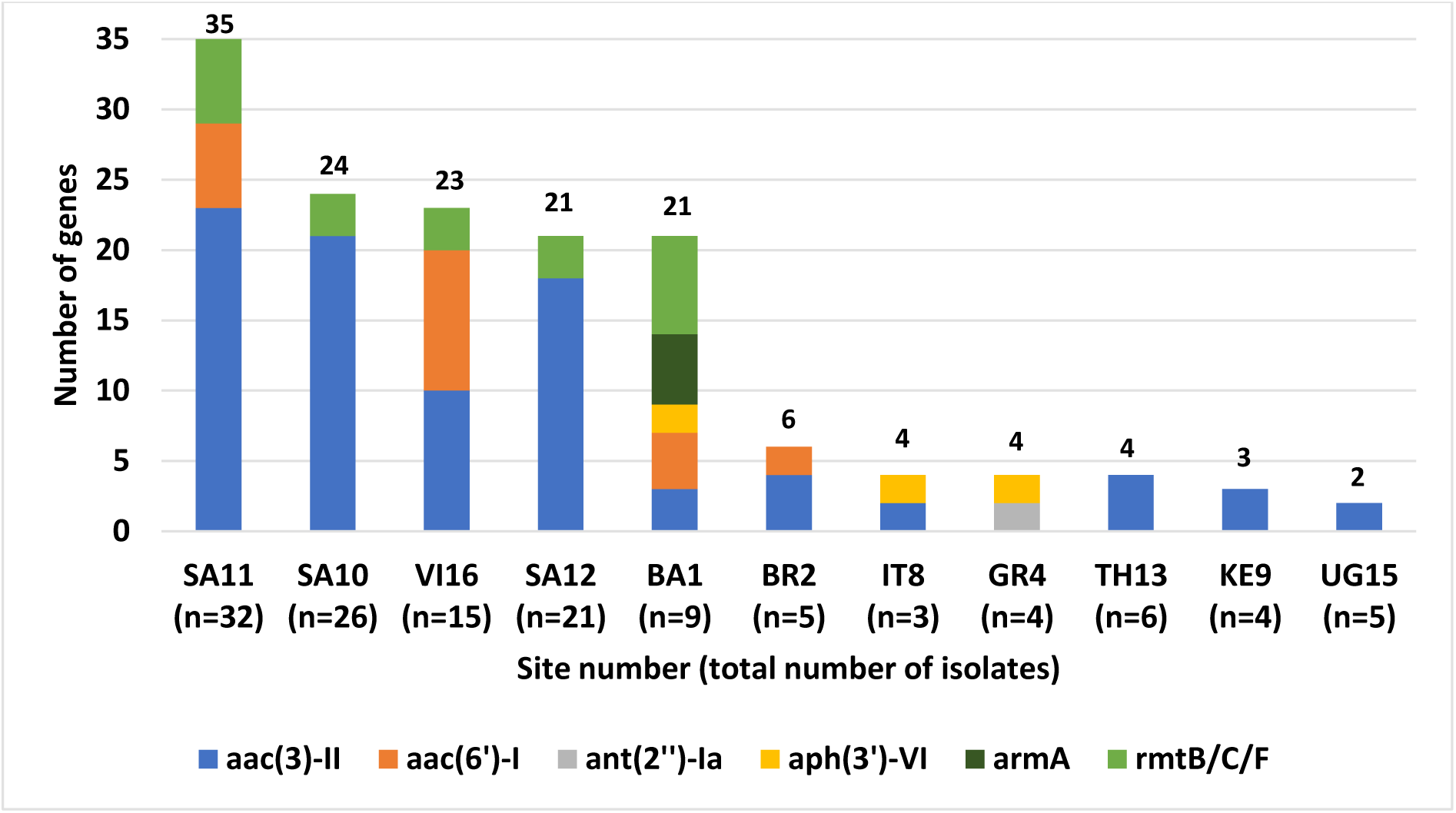
Distribution of aminoglycoside resistance genes of *K. pneumoniae* isolates (n=135) by site. (n) = total number of *K. pneumoniae* isolates per site found. Two sites that collected *K. pneumoniae* isolates (BR3 n=3 and TH14 n=2) but without aminoglycoside resistance genes being detected are not represented in this figure. Several isolates were carrying more than one resistance gene to a given class of antimicrobial agents. Other aminoglycoside resistance genes: *aadA, aph(6’)-I* (streptomycin resistance) and *aph(3’)-I* (neomycin/kanamycin resistance) were very frequent but are not represented in this figure due to their lack of clinical relevance in humans.

### Characterization of *E. coli*

Thirteen different *E. coli* STs were identified. ST1193 (n=13, 38%) was the most abundant in isolates from Vietnam (VI16, n=5) and one site in South Africa (SA11, n=4). The second most prevalent ST type was ST131 (n=5, 15%), reported from two sites in South Africa (SA10, n=1 and SA12, n=2), Thailand (TH13, n=1) and Vietnam (VI16, n=1).

Compared to *K. pneumoniae, E. coli* isolates carried fewer antibiotic resistance genes. Thirty-eight percent of the isolates (13/34) harbored an ESBL gene, mostly *bla*_CTX-M-27_ (n=8) and *bla*_CTX-M-15_ (n=5) genes. Only one isolate carried a *bla*_KPC-2_ carbapenem resistance gene (site VI16) and one with AmpC gene (*bla*_CMY-2_, site TH13).

Aminoglycoside resistance genes were detected in 9 *E. coli* isolates from 5 sites. Genes encoding AMEs modifying only gentamicin (*aac(3)-II* and *ant(2’)-Ia*) were observed in 7 isolates and were found in association with ESBL genes mostly in ST131 or in ST1193. Furthermore, two *E. coli* strains from site VI16 carried the *rmtB* 16S RNA methylase gene leading to resistance to all aminoglycosides including amikacin.

### Characterization of *A. baumannii*

In *A. baumannii*, 13 different sequence types were identified, with two of them predominant: ST1 (n=36) and ST2 (n=24). These two STs represent the two major international pandemic lineages, GC1 and GC2, respectively. Two sites from South Africa (SA10 and SA12) accounted for 80% of the *A. baumannii* isolates. Two different clusters could be delineated within ST1 and ST2 each (Figure 5). While the ST2 strains were grouped in two distinct clusters and differed between sites SA10 and SA12, the ST1 strains of these two sites were also grouped in two different clusters, but closely related to each other with a small allelic loci distance (Figure 5). Other sporadic STs not associated with pandemic lineages were found at single sites.

**Figure 5.**
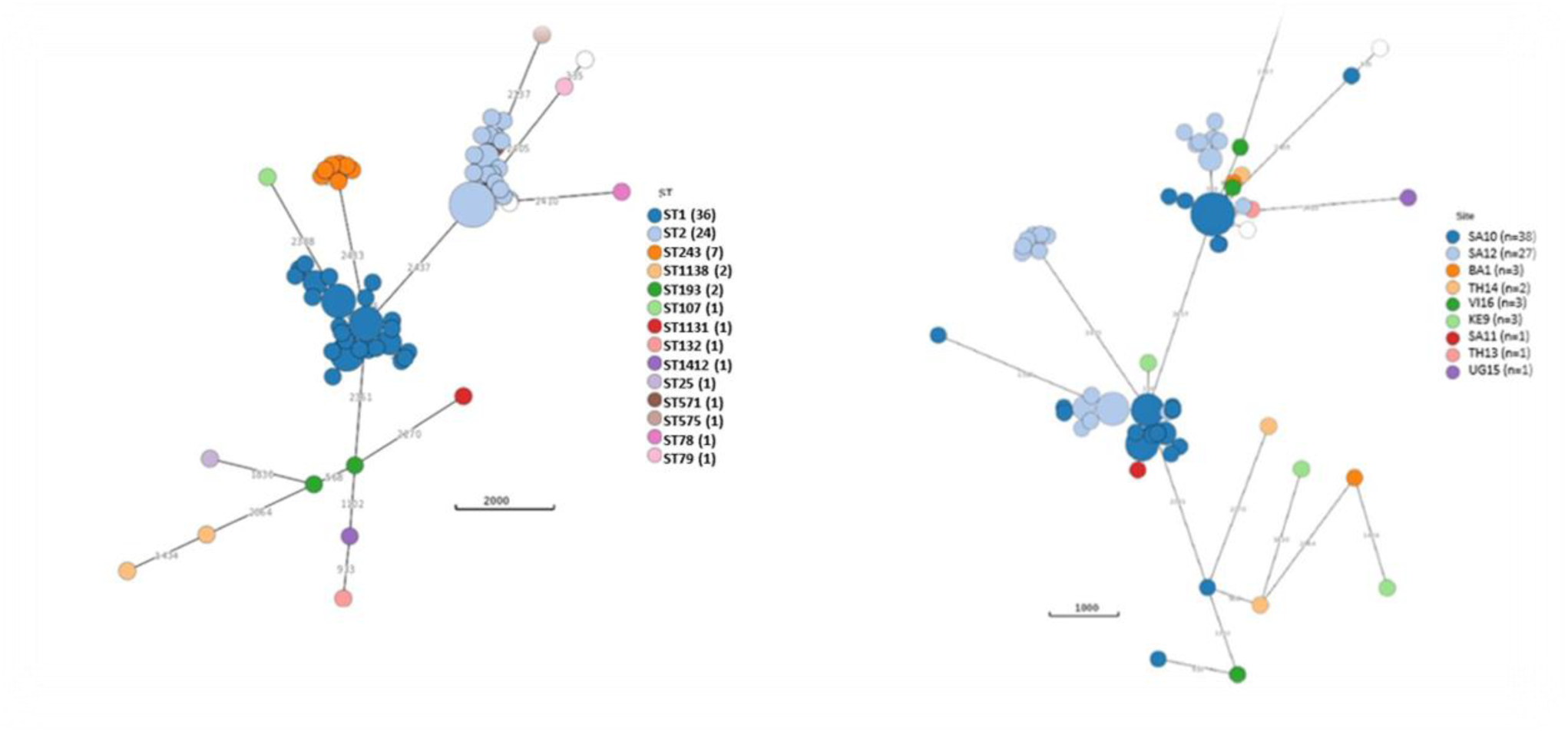
Minimum spanning tree from *A. baumannii* genomes (n=80) by cgMLST sequence types (STs). Left: The MLST groups (Pasteur scheme). Right: The sequence type diversity by site. White circles indicate the public references.

*A. baumannii* isolates displayed a very extensive drug resistance profile. Carbapenemase-producing *A. baumannii* isolates were found in 8 sites in 5 countries in Asia and Africa (Figure 6). *Bla_O_*_XA23_ was the most frequently present carbapenem resistance gene, present in 64/80 (80%) of the isolates and nearly half of the OXA-23-producers (n=36; 45%) co-carried *bla*_NDM-1_. Ten isolates (13%), seven of which from site SA12, were carrying a *bla*_OXA-58_ gene (Figure 6). All ST1 isolates from sites SA10, SA11 and SA12 in South Africa always carried the *bla*_NDM-1_ and *bla*_OXA-23_ genes. On the other hand, strains belonging to ST2 carried *bla*_OXA-23_ alone and never in association with *bla*_NDM-1_. Most of the *bla*_OXA-58_ positive isolates (n=10) were associated with ST243 and were found almost exclusively at site SA12 (n=7; 70%).

**Figure 6.**
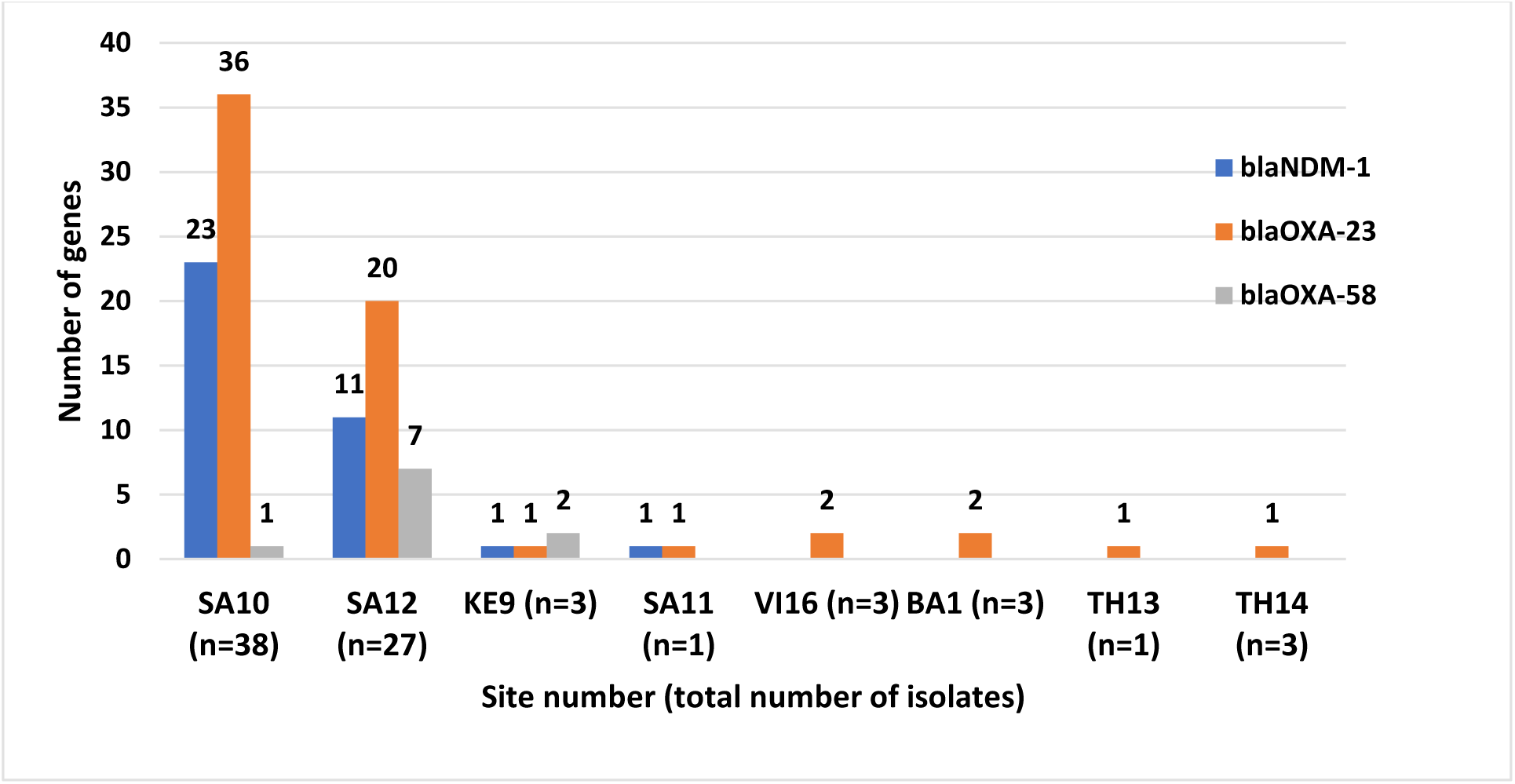
Distribution of carbapenem resistance genes in *A. baumannii* isolates (n=80) by site. Site UG15 (n=1) not represented in the chart since one unique *A. baumannii* isolate lacking any acquired carbapenemase producing genes.

More than 90% (74/80) of the *A. baumannii* isolates harbored one or several resistance mechanisms to all aminoglycosides including amikacin (Figure 7). The *armA* rRNA 16S methylase gene (in 53/80 strains, 62%) often in association with *aac(3)-Ia* (in 35/80 strains, 44%), was the most common aminoglycoside resistance mechanism.

**Figure 7.**
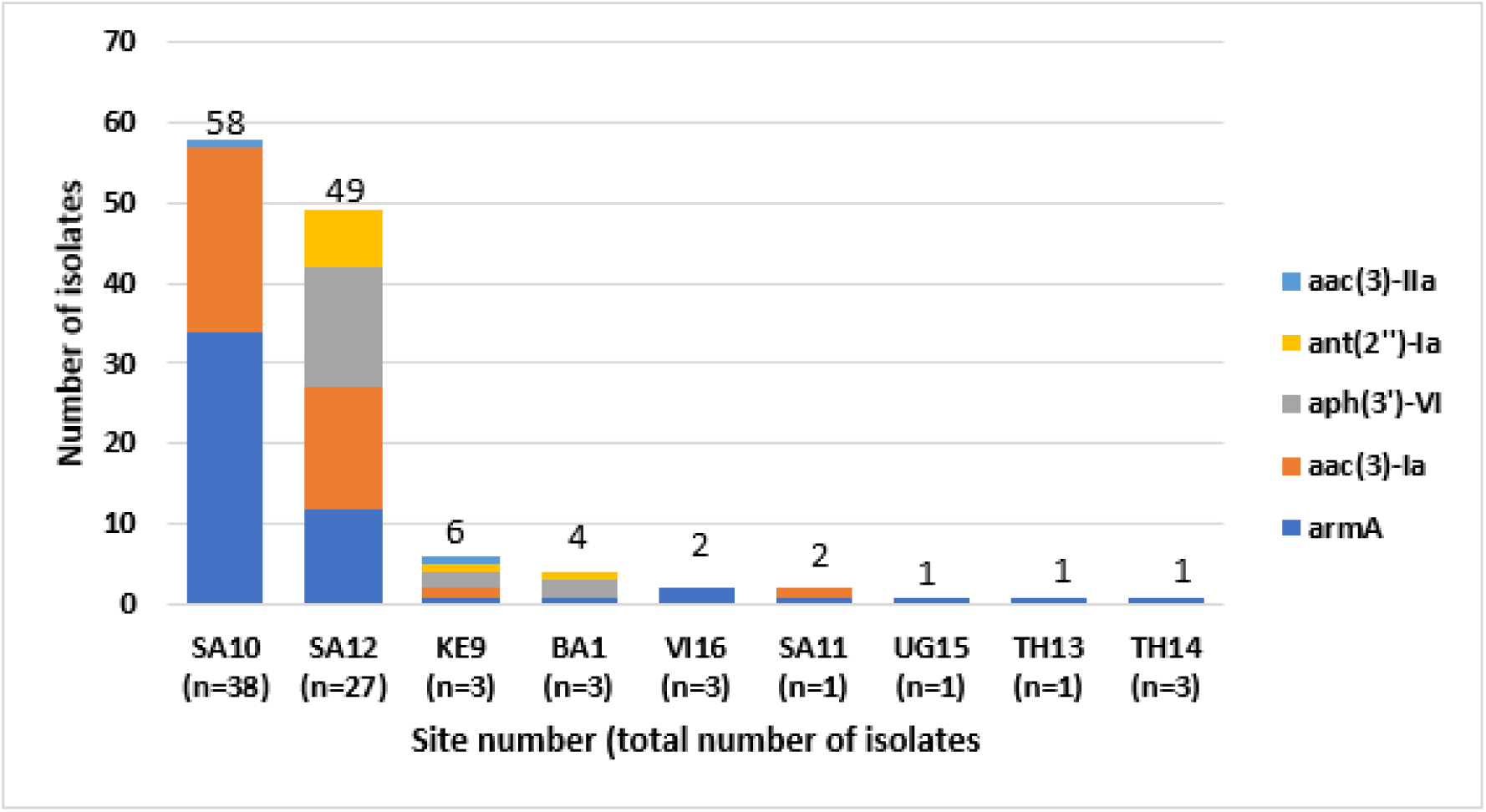
Distribution of aminoglycoside resistance genes of *A. baumannii* isolates (n=80) per site. (n) = total number of *A. baumannii* isolates collected per site. *aph(3’)-VI* (n=2) and *aph(3’)-VIa* (n=17) grouped together. Several isolates were carrying more than one resistance gene to a given class of antimicrobial agents.

### Characterization of *Enterobacter cloacae* complex (ECC)

ECC isolates (n=27) were collected at 12 sites in 8 countries with different subspecies occurring at the different sites. *E. hormaechei* (n=15) was the most common species found at 8 sites; *E. roggenkampii* (n=4) at two and *E. asburiae* (n=3) at 3 sites. Antimicrobial resistance in ECC is partly due to intrinsic chromosomal genes. All strains had in their core genome different alleles of *bla*_ACT_, an intrinsic AmpC gene that when overexpressed leads to resistance to expanded spectrum cephalosporins.

Multiple acquired resistance genes were found in 10/27 (37%) of the ECC isolates. All of these multidrug resistant strains were identified genetically as *E. hormaechei*. These strains belong to five different ST types including the well-known ST68 and ST78 MDR lineages. Nine of the 10 ESBL-producing *E. hormaechei* isolates carried a *bla*_CTX-M-15_ and one a *bla*_SHV-12_ gene. Other resistance genes that were frequently found in association with ESBLs were *bla*_OXA-1_ gene (resistance to piperacillin-tazobactam) and *aac(3)-II* coding genes (resistance to gentamicin). One of the 10 strains collected at site BA1 was also carrying a *bla*_NDM-1_ gene and a *rmtB* 16S rRNA methylase gene rendering it almost pan-resistant.

### Characterization of *Serratia marcescens*

Thirty-three *Serratia marcescens* isolates were collected from 7 sites in 6 countries, majority (n=19; 58%) from the two South African sites. The cgMLST analysis highlighted the diversity of the *S. marcescens* isolates, between the different sites but also within single sites.

Genes coding for ESBLs, *bla*_VEB-5_ (n=2), *bla*_CTX-M-14_ (n=2) and *bla*_CTX-M-15_ (n=2) were found in 6 (18%) isolates in Vietnam (n=3), Bangladesh (n=2), and Thailand (n=1). One isolate from Vietnam harbored a *bla*_NDM-5_ carbapenem resistance gene.

16S rRNA methylase genes (*armA* (n=2); *rmtB* (n=1)) were found in three isolates at two sites, BA1 and VI16.

## Discussion

Three generic antibiotics, flomoxef, amikacin and fosfomycin, were evaluated in novel combinations as new empiric carbapenem-sparing regimens for the treatment of neonatal sepsis, exhibited very good activity against the majority of these Enterobacterales isolates. The NeoOBS clinical study documented Gram-negative bacteria as the most common causative pathogens in neonatal sepsis [6], which is consistent with recent literature in LMICs [33]. The most frequently isolated species were *K. pneumoniae* and *A. baumannii,* a finding which is also in line with data by others from several single and multicenter studies of neonatal sepsis in LMICs [4, 5, 7, 9, 34–36]. *A. baumannii* isolates were predicted to be extremely drug-resistant as they harbored a large array of genes conferring resistance to almost all classes of antibiotics, leaving colistin as the only therapeutic option for neonatal sepsis caused by this organism. Among Enterobacterales, resistance to β-lactams was due to the widespread distribution of ESBLs, especially among *K. pneumoniae* (79%) and *E. coli* (38%) isolates. Piperacillin-tazobactam, that provides partial ESBL/pseudomonal coverage and was commonly used as empirical treatment by some hospitals [6], had moderate antibacterial activity against *E. coli* (susceptibility of 67%) but low activity against *K. pneumoniae* (susceptibility of 35% only), due to the widespread distribution of *bla*_OXA-1_. These high rates of antimicrobial resistance have been translated to the increased use of meropenem for treatment of these patients with sepsis at these sites. However, in this study resistance to meropenem was also observed in approximately 30% of *K. pneumoniae* isolates.

All isolates in this study showed wide genetic diversity, especially *K. pneumoniae* isolates, with 56 different STs in 135 isolates. Multidrug resistant strains belonged to different lineages and clones. Most of these isolates belong to multidrug-resistant international lineages that are already widely reported in both adults and neonates, and are known to be associated with nosocomial outbreaks and endemic hospital settings [37–39]. Besides their ability to spread through clonal expansion, these MDR lineages are also known to carry several plasmids, transposons and other mobile genetic elements that allow them to acquire and spread antimicrobial resistance within and across different bacterial species [40].

In *E. coli*, ST131 and ST1193 accounted as the most frequent STs in the NeoOBS study. This is in line with a recent epidemiological study which found that next to ST131, ST1993 is the second most widely globally distributed MDR clone recorded to date [41]. Interestingly, we found that *E. coli* ST1193 was also largely distributed in different sites from Vietnam to Kenya, Uganda and South Africa, highlighting the expansion of this lineage also on the African continent.

Multi-drug resistant *A. baumannii* strains were unevenly distributed at 8 sites, but found in large numbers at only two of the participating sites (SA10 and SA12).

*A. baumannii* has emerged during the last decade as a major difficult to treat nosocomial bacterial pathogen which is often linked with outbreaks or endemic contexts in healthcare settings. Once established, this organism can persist in the healthcare environment and is extremely difficult to eradicate [42]. Unlike *K. pneumoniae* and *E. coli* which showed high genetic diversity, *A. baumannii* isolates were less diverse and mostly belonged to the two dominant pandemic international clonal groups (CG1 and CG2) known to occur in a limited number of sites, mostly in South Africa.

Susceptibility of *S. marcescens* and ECC was inferred from genotyping of resistance genes. Both species had inducible chromosomal AmpC genes and showed resistance to the WHO second-line cephalosporin regimen (cefotaxime/ceftriaxone) as well as to flomoxef alone. It is well known from the literature that derepressed mutants can be selected in AmpC inducible species during therapy with these agents and lead to the development of resistance and clinical failure, especially in invasive infection caused by *Enterobacter cloacae* complex [43].

Although the study recruited over 3200 neonates, the limitations of this study mainly relate to the relatively small number of samples obtained, especially from sites where lower number of patients were enrolled. Besides differences in the prevalence and distribution of neonatal sepsis pathogens, variation in blood culture positivity rates between centers was also observed [6], possibly indicating differences in the collection and/or performance of microbiological methods used locally. The majority of neonatal units participating in this study were from tertiary hospitals in urban areas, so the burden of AMR may not be representative for district hospitals. These elements represent an important bias in most AMR studies in low-resource settings, where the need for high-quality microbiology means that certain settings may be overrepresented.

## Conclusion

The NeoOBS study showed wide variety of bacterial species as a cause of neonatal sepsis, many of which carried multiple resistance genes. Among Gram-negative isolates, *K. pneumoniae* was the most predominant species widely distributed among the sites. Many of the isolates from our study were resistant to the first and second line WHO treatments, mostly because of the very frequent occurrence of ESBLs. New agents that can target these pathogens are urgently needed. The three novel antibiotic combinations, fosfomycin-flomoxef, fosfomycin-amikacin and flomoxef-fosfomycin showed high activity especially against ESBL producing *E. coli* and *Klebsiella pneumoniae* isolates. Our study supports the evaluation of these combinations in the NeoSep1 trial, for treatment of neonatal sepsis.

## Data Availability

All data produced in the present study are available upon reasonable request to the authors

