## Supplemental Table 1-3 for "Identification of potential novel combination antibiotic regimens based on drug-susceptibility and genetic diversity of Gram-negative bacteria causing neonatal sepsis in low- and middle-income countries"

**Supplementary Table 1. List of sites whose isolates were characterized at University of Antwerp (UA).**

| **Country** | **UA Site Number** | **Country/site number codes** |
| --- | --- | --- |
| Bangladesh | Site 1 | BA1 |
| Brazil | Site 2 | BR2 |
| Brazil | Site 3 | BR3 |
| Greece | Site 4 | GR4 |
| Italy | Site 8 | IT8 |
| Kenya | Site 9 | KE9 |
| South Africa | Site 10 | SA10 |
| South Africa | Site 11 | SA11 |
| South Africa | Site 12 | SA12 |
| Thailand | Site 13 | TH13 |
| Thailand | Site 14 | TH14 |
| Uganda | Site 15 | UG15 |
| Vietnam | Site 16 | VI16 |

**Supplementary Table 2. In vitro susceptibility of *K. pneumoniae* (n=87) *and E. coli* (n=21) isolates to selected antimicrobial agents.** MIC ranges, MIC_50_, MIC_90_ values and percentages of resistance are according to the EUCAST clinical breakpoints (v11.0, January 2021).

| **Antibiotic** | **MIC ranges** | **MIC_50_ (mg/L)** | **MIC_90_ (mg/L)** | **EUCAST R%** |
| --- | --- | --- | --- | --- |
| ***K. pneumoniae*** |  |  |  |  |
| Ampicillin | 8- >128 | >128 | >128 | 98 |
| Piperacillin-Tazobactam | 8/4- >256/4 | 32/4 | >256/4 | 65 |
| Flomoxef | 0.06- >128 | 0.25 | >128 | 34* |
| Cefotaxime | 0.03- >128 | >128 | >128 | 85 |
| Meropenem | 0.03- >128 | 0.06 | >128 | 30 |
| Amikacin | 1- >256 | 4 | >256 | 31 |
| Gentamicin | 0.25- >256 | 64 | >256 | 74 |
| Fosfomycin | 1- >512 | 8 | 64 | 11 |
| ***E. coli*** |  |  |  |  |
| Ampicillin | 4->128 | >128 | >128 | 90 |
| Piperacillin-Tazobactam | 1/4->256/4 | 2/4 | 32/4 | 33 |
| Flomoxef | <=0.06- >128 | 0.12 | 0.5 | 10* |
| Cefotaxime | 0.06->128 | 0,5 | >128 | 47 |
| Meropenem | 0.03-32 | 0.03 | 0.06 | 5 |
| Amikacin | 2->256 | 4 | 32 | 14 |
| Gentamicin | 0.5->256 | 2 | 256 | 38 |
| Fosfomycin | 0.5->512 | 0.5 | 1 | 5 |

*Isolates were reported as susceptible to flomoxef up to a MIC of ≤1 mg/L [34]

**Supplementary Table 3. Minimal inhibitory concentrations (MICs) for all *E. coli* and *K. pneumoniae* strains in NeoOBS study tested against six antibiotics and their susceptibility against the three new antibiotic combinations (Fosfomycin/flomoxef; Fosfomycin/amikacin and flomoxef/amikacin)**

|  | **Strain Code** | **AMP/GEN** | **TZP** | **CTX** | **MEM** | **FLX** | **AMK** | **FOS** | **FOS/FLX** | **FOS/AMK** | **FLX/AMK** |
| --- | --- | --- | --- | --- | --- | --- | --- | --- | --- | --- | --- |
| *K. pneumoniae* | MIC00063530 | 64 | 16/4 | >128 | 0.06 | 0.25 | 2 | 16 | S | S | S |
|  | MIC00063533 | 32 | 32/4 | >128 | 0.06 | 0.12 | 4 | 32 | S | S | S |
|  | MIC00063560 | 0,5 | 4/4 | >128 | 0.12 | 0.12 | 1 | 16 | S | S | S |
|  | MIC00063612 | 64 | 32/4 | >128 | 0.06 | 0.25 | 4 | 8 | S | S | S |
|  | MIC00063641 | 2 | 2/4 | 0,062 | 0.03 | <=0.06 | 2 | 8 | S | S | S |
|  | MIC00063649 | 1 | 4/4 | 0,031 | 0.03 | <=0.06 | 1 | 8 | S | S | S |
|  | MIC00063650 | 16 | 64/4 | 64 | 0.03 | 0.12 | 8 | 8 | S | S | S |
|  | MIC00063653 | >256 | >256/4 | >128 | >128 | >128 | >256 | 8 | R | R | R |
|  | MIC00063920 | >256 | >256/4 | >128 | >128 | >128 | >256 | 8 | R | R | R |
|  | MIC00063923 | 16 | 16/4 | >128 | 0.06 | 1 | 2 | 64 | S | S | S |
|  | MIC00063927 | 64 | 16/4 | >128 | 0.06 | 0.12 | 8 | 16 | S | S | S |
|  | MIC00063932 | 16 | >256/4 | >128 | 16 | >128 | 8 | 16 | R | S | R |
|  | MIC00063953 | 32 | 16/4 | >128 | 0.06 | 0.12 | 8 | 4 | S | S | S |
|  | MIC00063954 | 64 | 8/4 | >128 | 0.03 | 0.12 | 4 | 4 | S | S | S |
|  | MIC00063959 | 64 | 256/4 | >128 | 0.06 | 0.12 | 4 | 4 | S | S | S |
|  | MIC00065380 | >256 | 16/4 | >128 | 0.12 | 0.12 | >256 | 4 | S | R | S |
|  | MIC00065382 | >256 | >256/4 | >128 | 128 | >128 | >256 | 8 | R | R | R |
|  | MIC00065387 | 128 | 4/4 | >128 | 0.06 | 0.12 | 2 | 64 | S | S | S |
|  | MIC00065403 | 128 | >256/4 | >128 | >128 | >128 | 16 | 8 | R | S | R |
|  | MIC00065410 | 64 | >256/4 | 4 | 0.25 | 128 | 2 | 8 | R | S | R |
|  | MIC00065413 | 0,5 | 64/4 | >128 | 0.06 | 0.25 | 2 | 16 | S | S | S |
|  | MIC00065418 | 4 | >256/4 | >128 | 128 | 64 | 8 | 8 | S | S | R |
|  | MIC00065422 | 128 | 256/4 | >128 | 32 | 64 | 16 | 2 | S | S | R |
|  | MIC00065435 | 128 | >256/4 | >128 | >128 | >128 | 16 | 8 | R | S | R |
|  | MIC00065437 | 128 | >256/4 | >128 | >128 | >128 | 16 | 8 | R | S | R |
|  | MIC00065438 | 256 | >256/4 | >128 | >128 | >128 | 16 | 4 | R | S | R |
|  | MIC00065588 | >256 | >256/4 | >128 | 128 | 128 | >256 | 16 | R | R | R |
|  | MIC00065592 | 128 | >256/4 | >128 | >128 | >128 | 16 | 8 | R | S | R |
|  | MIC00065700 | 32 | 64/4 | >128 | 0.06 | 0.12 | 8 | 32 | S | S | S |
|  | MIC00065711 | 64 | 16/4 | >128 | 0.06 | 0.12 | 8 | 32 | S | S | S |
|  | MIC00065745 | 0,5 | 4/4 | 0,031 | 0.03 | 0.06 | 2 | 8 | S | S | S |
|  | MIC00065748 | 16 | 8/4 | >128 | 0.06 | 0.12 | 8 | 8 | S | S | S |
|  | MIC00065749 | 32 | 32/4 | >128 | 0.06 | 0.12 | 4 | 8 | S | S | S |
|  | MIC00065750 | >256 | >256/4 | >128 | 64 | 128 | >256 | 16 | R | R | R |
|  | MIC00065755 | 32 | 4/4 | >128 | 0.06 | 0.12 | 2 | 16 | S | S | S |
|  | MIC00065772 | 1 | >256/4 | >128 | 1 | 0.5 | 4 | 16 | S | S | S |
|  | MIC00065779 | >256 | 32/4 | >128 | 0.06 | 0.12 | 16 | 32 | S | R | S |
|  | MIC00065788 | 128 | 256/4 | >128 | 0.06 | 0.12 | 8 | 64 | S | R | S |
|  | MIC00065795 | 64 | 8/4 | >128 | 0.06 | 0.12 | 2 | 32 | S | S | S |
|  | MIC00065796 | 64 | 4/4 | >128 | 0.06 | 0.12 | 2 | 8 | S | S | S |
|  | MIC00065802 | 1 | 4/4 | 0,125 | 0.06 | 0.12 | 2 | >512 | S | R | S |
|  | MIC00065806 | 1 | 256/4 | >128 | 0.06 | 0.25 | 2 | 8 | S | S | S |
|  | MIC00065808 | 256 | 32/4 | >128 | 0.12 | 0.25 | 4 | 8 | S | S | S |
|  | MIC00065814 | 64 | 16/4 | >128 | 0.06 | 0.12 | 4 | 8 | S | S | S |
|  | MIC00065821 | >256 | >256/4 | >128 | 64 | 128 | >256 | 128 | R | R | R |
|  | MIC00065828 | >256 | >256/4 | >128 | >128 | >128 | >256 | 8 | R | R | R |
|  | MIC00065830 | 1 | >256/4 | >128 | 128 | >128 | 32 | 16 | R | R | R |
|  | MIC00065835 | >256 | >256/4 | >128 | 4 | 4 | >256 | 16 | S | R | R |
|  | MIC00065836 | >256 | >256/4 | >128 | 128 | >128 | >256 | 8 | R | R | R |
|  | MIC00065844 | 0,5 | 8/4 | >128 | 0.12 | 0.12 | 2 | 32 | S | S | S |
|  | MIC00065851 | >256 | >256/4 | >128 | 128 | >128 | >256 | 16 | R | R | R |
|  | MIC00065854 | >256 | >256/4 | >128 | 128 | >128 | >256 | 8 | R | R | R |
|  | MIC00065858 | >256 | >256/4 | >128 | >128 | >128 | >256 | 128 | R | R | R |
|  | MIC00065961 | 0,25 | 4/4 | 16 | 0.06 | 16 | 1 | 64 | R | S | S |
|  | MIC00065973 | 16 | 8/4 | 4 | 0.06 | 32 | 256 | 16 | S | R | R |
|  | MIC00065974 | 16 | 8/4 | 16 | 0.06 | 32 | 128 | 8 | S | R | R |
|  | MIC00065988 | 0,25 | 4/4 | 0,062 | 0.06 | 0.12 | 1 | 4 | S | S | S |
|  | MIC00066061 | 0,5 | 8/4 | >128 | 0.06 | 0.12 | 2 | 4 | S | S | S |
|  | MIC00066090 | 0,5 | 4/4 | 0,062 | 0.03 | 0.12 | 2 | 8 | S | S | S |
|  | MIC00066112 | 128 | >256/4 | >128 | 0.06 | 0.25 | 8 | 4 | S | S | S |
|  | MIC00066124 | 128 | 256/4 | >128 | 0.06 | 0.25 | 4 | 8 | S | S | S |
|  | MIC00066136 | 128 | 32/4 | >128 | 0.06 | 0.25 | 2 | 16 | S | S | S |
|  | MIC00066154 | 64 | 16/4 | >128 | 0.06 | 0.25 | 4 | 8 | S | S | S |
|  | MIC00066174 | 64 | 16/4 | >128 | 0.06 | 0.12 | 8 | 2 | S | S | S |
|  | MIC00066218 | 16 | 4/4 | >128 | 0.06 | 0.12 | 2 | 16 | S | S | S |
|  | MIC00066226 | 128 | 128/4 | >128 | 0.06 | 0.25 | 4 | 8 | S | S | S |
|  | MIC00066233 | 64 | 32/4 | >128 | 0.06 | 0.25 | 4 | 16 | S | S | S |
|  | MIC00066260 | 0,5 | 2/4 | 0,062 | 006 | <=0.06 | 2 | 8 | S | S | S |
|  | MIC00066263 | 64 | 2/4 | 64 | 0.03 | 0.12 | 4 | 8 | S | S | S |
|  | MIC00066276 | 32 | >256/4 | >128 | 128 | >128 | 4 | 4 | R | S | R |
|  | MIC00066289 | 0,25 | 128/4 | 64 | 1 | 128 | 2 | 16 | R | S | R |
|  | MIC00066304 | 128 | 256/4 | >128 | 0.06 | 0.25 | 2 | 1 | S | S | S |
|  | MIC00066328 | 64 | 4/4 | 0,5 | 0.03 | 0.12 | 16 | 8 | S | S | S |
|  | MIC00066364 | 0,5 | 4/4 | 0,125 | 0.06 | 0.25 | 2 | 16 | S | S | S |
|  | MIC00066387 | 0,5 | 2/4 | 0,062 | 0.06 | 0.12 | 2 | 2 | S | S | S |
|  | MIC00066439 | 128 | >256/4 | >128 | 64 | >128 | 4 | 16 | R | S | R |
|  | MIC00066443 | 256 | >256/4 | >128 | >128 | >128 | 16 | 128 | R | R | R |
|  | MIC00066448 | 0,5 | 4/4 | 0,25 | 0.06 | 0.25 | 1 | 64 | S | S | S |
|  | MIC00066461 | 256 | >256/4 | 64 | >128 | >128 | 16 | 64 | R | R | R |
|  | MIC00066466 | 0,5 | 4/4 | 0,062 | 0.06 | 0.12 | 1 | 8 | S | S | S |
|  | MIC00066469 | 0,5 | 2/4 | 0,062 | 0.06 | 0.12 | 1 | 16 | S | S | S |
|  | MIC00066476 | 128 | >256/4 | >128 | 0.25 | 0.25 | 2 | 4 | S | S | S |
|  | MIC00066484 | 0,5 | 8/4 | >128 | 0.06 | 0.25 | 4 | 4 | S | S | S |
|  | MIC00066513 | 64 | 8/4 | >128 | 0.06 | 0.12 | 1 | 8 | S | S | S |
|  | MIC00066515 | 0,5 | 4/4 | >128 | 0.06 | 0.12 | 1 | 8 | S | S | S |
|  | MIC00066518 | 64 | 32/4 | >128 | 0.12 | 0.12 | 1 | 8 | S | S | S |
|  | MIC00066523 | 64 | 16/4 | >128 | 0.06 | 0.12 | 16 | 4 | S | S | S |
| *E. coli* | MIC00063566 | 1 | 2/4 | 0,062 | 0.03 | <=0.06 | 2 | 0.5 | S | S | S |
|  | MIC00063571 | 256 | 16/4 | >128 | 0.03 | 0.5 | 8 | 1 | S | S | S |
|  | MIC00063935 | 2 | 2/4 | >128 | 0.03 | 0.12 | 4 | 1 | S | S | S |
|  | MIC00065379 | 1 | ¼ | >128 | 0.03 | 0.12 | 2 | 0.5 | S | S | S |
|  | MIC00065390 | >256 | 2/4 | >128 | 0.03 | 0.12 | >256 | 4 | S | R | S |
|  | MIC00065415 | 0,5 | 32/4 | 0,5 | 0.03 | 0.12 | 4 | 0.5 | S | S | S |
|  | MIC00065427 | >256 | >256/4 | >128 | 32 | >128 | >256 | >512 | R | R | R |
|  | MIC00065710 | 0,5 | 4/4 | 0,125 | 0.03 | 0.12 | 2 | 1 | S | S | S |
|  | MIC00065719 | 2 | 2/4 | 0,062 | 0.03 | <=0.06 | 2 | 0.5 | S | S | S |
|  | MIC00065722 | 2 | 2/4 | 0,062 | 0.03 | 0.12 | 8 | 0.5 | S | S | S |
|  | MIC00065833 | 4 | 2/4 | >128 | 0.03 | 0.12 | 16 | 0.5 | S | S | S |
|  | MIC00065964 | 1 | 2/4 | 0,062 | 0.03 | <=0.06 | 4 | 0.5 | S | S | S |
|  | MIC00065983 | 2 | 16/4 | 0,25 | 0.03 | <=0.06 | 4 | 0.5 | S | S | S |
|  | MIC00066023 | 32 | ¼ | 0,062 | 0.03 | 0.12 | 4 | 1 | S | S | S |
|  | MIC00066074 | 8 | 16/4 | 0,25 | 0.03 | 0.12 | 4 | 1 | S | S | S |
|  | MIC00066198 | 4 | 2/4 | >128 | 0.03 | 0.12 | 8 | 0.5 | S | S | S |
|  | MIC00066203 | 1 | 2/4 | 0,125 | 0.03 | 0.12 | 4 | 0.5 | S | S | S |
|  | MIC00066275 | 64 | 4/4 | 8 | 0.06 | 64 | 4 | 0.5 | S | S | R |
|  | MIC00066287 | 128 | 64/4 | >128 | 0.06 | 0.5 | 16 | 0.5 | S | S | S |
|  | MIC00066329 | 2 | 2/4 | 0,125 | 0.03 | 0.12 | 4 | 0.5 | S | S | S |
|  | MIC00066468 | 64 | 16/4 | >128 | 0.06 | 0.12 | 32 | 0.5 | S | S | S |
|  | **Coverage (%)** | **32** | **41** | **22** | **76** | **70** | **70** | **90** | **76** | **79** | **71** |
